# Elevated Neutrophil to Lymphocyte Ratio Predicts In-hospital Mortality Among Stroke Patients in a Metropolitan hospital in Australia, Universal Value-added measure in Stroke Care

**DOI:** 10.1101/2021.03.01.21252317

**Authors:** Tissa Wijeratne, Carmela Sales, Leila Karimi, Mihajlo Jakovljevic

## Abstract

Neutrophil counts (innate immunity) and lymphocyte counts (adaptive immunity) are common markers of inflammation in the context of acute stroke, and Neutrophil lymphocyte ratio (NLR) is likely to be expected to predict the post-stroke outcome.

This study aimed to explore the predictive value of NLR to predict the very early mortality during the acute hospital admission (death within the first week of hospital stay) as this has management implications for the ongoing investigations, family discussions and resource allocation. This the first such study attempting at exploring the role of NLR in hyperacute in-patient mortality in the world to the best of our knowledge.

This retrospective study included 120 patients (60 acute stroke patients who died within the first week of the hospital stay and 60 age, sex-matched controls who were discharged within two weeks of the hospital admission, alive. We reviewed the total white cell counts of these patients (first 72 hours of the hospital admission) and NLR was calculated manually. While there was no statistically significant difference between mean neutrophil counts and mean lymphocyte counts between the two cohorts [neutrophil counts (mean, SD), 8.52(3.20) in the death cohort, 6.48(2.20) among survivors and lymphocyte (mean, SD), 1.83(1.48) in the death cohort,1.66 (0.69) among survivors], there was a statistically significant difference in NLR between the two cohorts. NLR (mean, SD) was 6.51 (4.98) in the death cohort while the survivors had mean NLR of 4.64 with an SD 2.90 with a p-value of 0.048. Hypertension, diabetes, atrial fibrillation, previous vascular events were common in both groups indicating the value of exploring the evidence of background compromised vascular system and metabolic syndrome and bringing the systems biology approach to the management of stroke.

## Introduction

Stroke is a devastating condition which results in significant mortality(1, 2) and morbidity despite advancement in thrombolytic therapy and endovascular interventions and general improvement in healthcare. COVID-19 and acute stroke appear to share common pathobiological mechanisms with a renewed interest in NLR and immune-based biomarkers in both these fields, making this aspect a priority for the front-line clinicians(3).

Besides it exposes substantial capability to influence fiscal sustainability(4) of health care budget allocated for noncommunicable diseases *(NCDs) even among richest societies(5, 6). This phenomenon appears to be even more prominent and challenging among the Emerging Markets such as the BRICS(7) and EM7(8) which are now days shaping entire global demand and supply for stroke – related medical goods and services. Various factors are known to impact in-hospital mortality, including severe stroke at presentation, age, gender and topographic location (9). Biomarkers have also been studied to identify stroke outcomes in the short and long term.

The neutrophil to lymphocyte ratio (NLR) has been shown to predict clinical outcomes in ischemic and haemorrhagic stroke (10-19). Yu and colleagues demonstrated that patients with higher NLR values on admission have more severe neurologic dysfunction at discharge (14). It has also been shown that NLR among patients with intracerebral haemorrhage likewise predicted mortality in the short term. A meta-analysis of 37 studies confirms that NLR’s utility in ascertaining prognosis at three months in ischemic and haemorrhagic strokes supports the overwhelming literature in significance in innate and adaptive immune activity in acute stroke and the recovery process(20-23).

While NLR has been utilized in various aspects of stroke prognostication, little is known about its use in predicting in-hospital mortality, especially in the acute stage. This study investigates the differences in NLR among stroke deaths and survivors within the said period. It also aims to determine the specificity and sensitivity in predicting mortality in this population. We believe the systems biology-based approach in acute stroke care is the key to further advances in stroke medicine.

## Method

This study retrospectively analyzed prospectively obtained clinical records of stroke patients admitted at Western Health from January 1 to December 31, 2017, at Western Health, Melbourne Australia as part of the stroke thrombolysis registry. The local institutional review board approved the study.

This study included 120 strokes (ischemic or intracerebral haemorrhage) patients who died within the first week of admission and were compared to age and sex-matched controls who were discharged within 14 days from admission. Patients were included if they were (1) >18 years old, (2) clinical and radiologic diagnosis of stroke, and (3) death of any cause documented within seven days form admission. Patients were excluded if they have (1) clinical evidence of systemic infection within 48 hours after treatment (2) known autoimmune disease, cancer and steroid therapy.

### Clinical protocol

Records of clinical and laboratory data were reviewed and documented using a pre-conceived data collection form. In particular, demographics, medical history, and comorbidities and concomitant medications were looked into. Documentation of stroke severity, neuroimaging, clinical course and outcomes were likewise investigated.

### Venous blood samples

Records of venous blood samples were reviewed. Bloods taken on admission and up to 72 hours from admission were of interest. Total leukocyte, neutrophil, and lymphocyte counts were determined using Sysmex® XN-Series10 by the nationally accredited hospital pathology provider as part of the routine assessment, and The NLR was calculated manually as the ratio of the percentage of neutrophils over the percentage of lymphocytes by TW with the help of excel software.

### Statistical analysis

Data analysis was performed using SPSS v.27. Categorical variables were expressed as frequency and percentages, while continuous variables were described as median and mean ± SD. Statistical significance was determined using the Pearson χ^2^ test and Kruskal-Wallis test for categorical and continuous variables, respectively. Receiver operating curve (ROC) was used to determine the predictive value of NLR as a predictor of in-hospital mortality among stroke patients. A p-value of LJ<LJ.05 was interpreted to be of statistically significant difference.

## Results

Table 1 shows the baseline characteristics of patients included in the study. There are no significant differences between groups in terms of age, sex, comorbidities and laboratory results. Statistical differences were noted in terms of baseline MRS and GCS, wherein patients who died had higher values for both on admission. Similarly, laboratory parameters such as mean neutrophil counts and NLR values were also significantly higher in the mortality group.

**Table 1.**

|  | Death<br>N=60 | Survived<br>N=60 | p-value |
| --- | --- | --- | --- |
| Age (years) | 79.2666667 | 78.98333333 | 0.84 |
| Sex (% female) | 56.6% (34) | 56.6% (34) | 1.00 |
| Modified Rankin Scale | 2.11666667 | 1.216666667 | 0.003 |
| History of stroke | 30% (24) | 43% (26) | 0.131 |
| Hypertension | 81.60% (49) | 83.30% (50) | 0.811 |
| Diabetes | 31.60% (20) | 30% (18) | 0.844 |
| Atrial Fibrillation | 31.60% (20) | 43.30% (26) | 0.189 |
| Chronic Kidney Disease | 10% (6) | 6.66% (40) | 0.511 |
| Dyslipidaemia | 46.60% (28) | 52% (31) | 0.585 |
| Current Antiplatelet | 35% (21) | 30% (18) | 0.560 |
| Current Statin | 36.60% (22) | 46.60% (28) | 0.269 |
| Previous/Active Smoking | 25% (15) | 33% (20) | 0.317 |
| Ischemic Stroke Aetiology | 66% (40) | 90% (54) | 0.002 |
| GCS (Median, range) | 10 (3-15) | 15 (9-15) | 0.000 |
| Systolic Blood Pressure<br>(Mean, SD) | 150.2(27.3) | 154.3 (32.47) | 0.544 |
| Diastolic Blood Pressure<br>(Mean, SD) | 83.6(16.35) | 82.4 (14.2) | 0.816 |
| Neutrophil (Mean, SD) | 8.52 (3.20) | 6.48 (2.20) | 0.010 |
| Lymphocyte (Mean, SD) | 1.83 (1.48) | 1.66 (0.69) | 0.476 |
| Neutrophil to Lymphocyte Ratio<br>(Mean, SD) | 6.51 (4.98) | 4.64 (2.90) | 0.048 |
| White Cell Count<br>(Mean, SD) | 11.04 (4.51) | 9.14 (3.30) | 0.13 |
| Haemoglobin, (Mean, SD) | 125.5(26.22) | 131.45(23.2) | 0.73 |
| Platelet (Mean, SD) | 258.21 (99) | 236.35 (62.75) | 0.407 |
| Platelet to lymphocyte ratio<br>(Mean, SD) | 184.73 (87.3) | 165.15 (82.11) | 0.99 |

**Figure 1a-1c.**
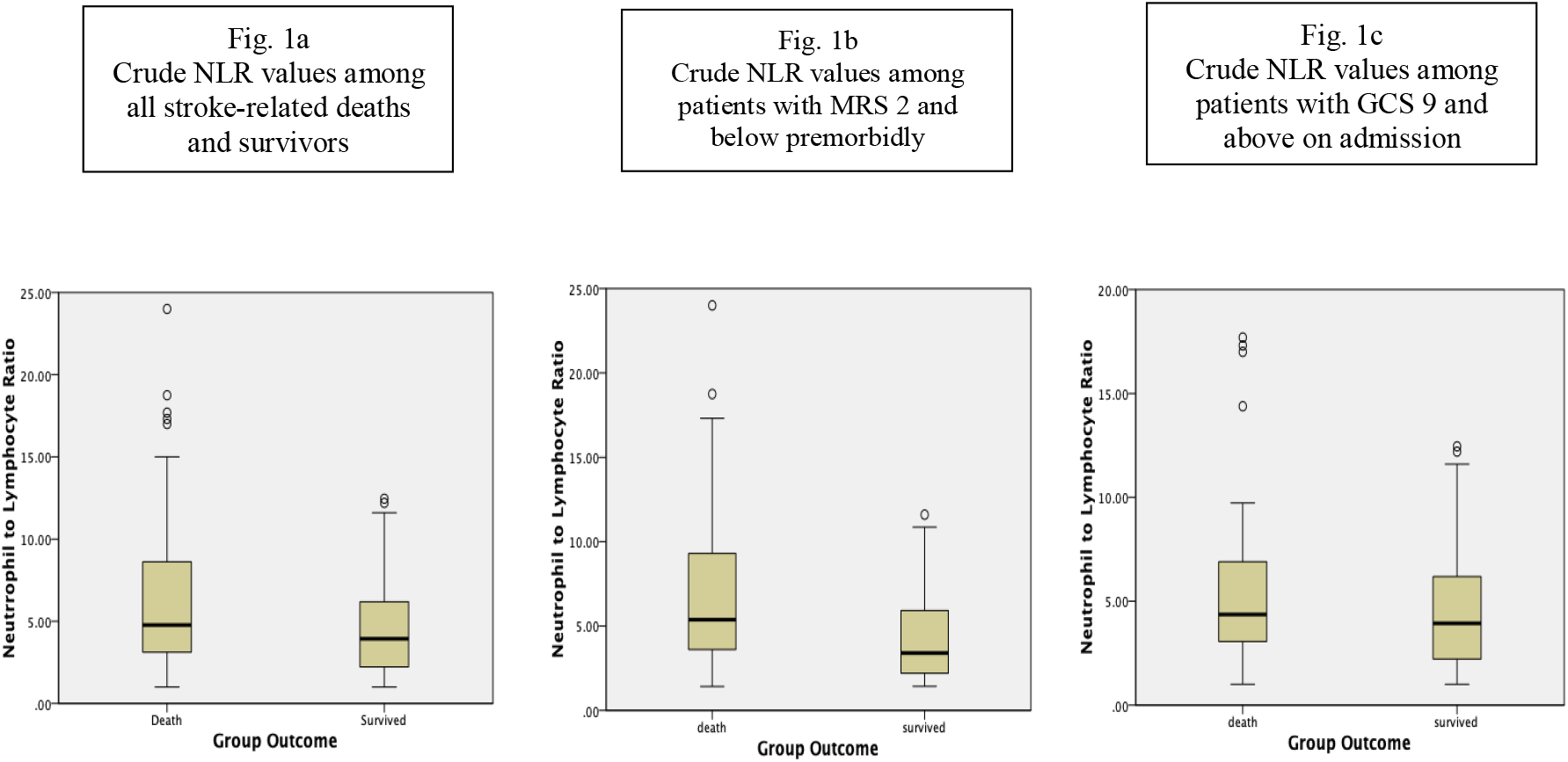
shows the NLR values of stroke-related deaths and mortalities in all patients, those with good functional status premorbid (mRS two and below) and patients with mild to moderate stroke (GCS 9 and above) at presentation. Statistically significant differences in NLR values were noted among all stroke-related deaths and survivors (p=0.048). Similar trends were observed when only patients with good functional premorbid were considered (p=0.010). Mean NLR values were also higher in the death group compared to the survivors when only patients with GCS 9 and above were considered, however, effects were not statistically significant (p=.227)

**Figure 2.**
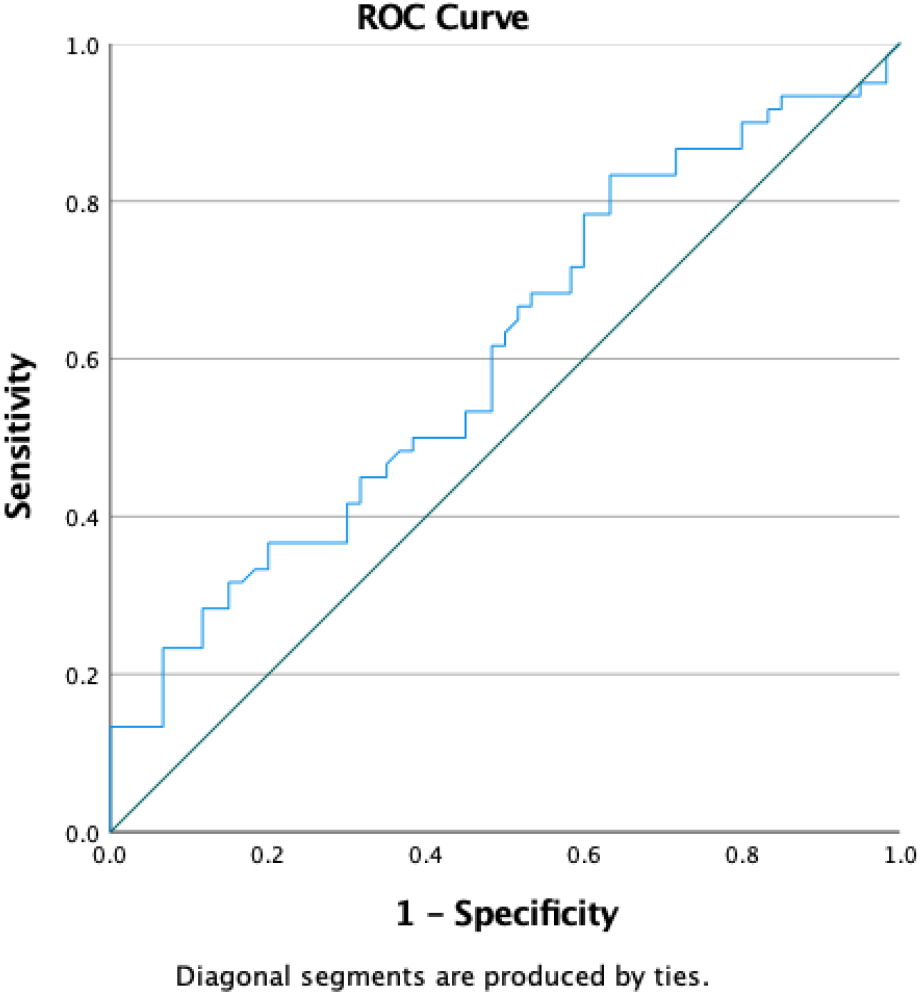
shows the Receiver Operator Curve (ROC) analysis utilizing NLR for predicting in-hospital mortality. Further analysis has been demonstrated that an NLR cut-off value of 1.54, an area under the curve of .605, yields a sensitivity of 93% and specificity of 90% and a p-value of 0.048 (95% CI 0.504-0.706).

## Discussion

The NLR is a useful predictor of in-hospital mortality in the acute period among stroke patients. Its prognosticating value may be limited because of its moderate sensitivity and specificity. Still, it may be useful in determining the risk of death among patients with good baseline functionality. The NLR is easily available anywhere in the world.

Inflammation is key to stroke’s pathogenesis, especially in the acute stage(24-30). It is known that neutrophils are the first cells to adhere to vessels within minutes following an acute vascular injury which triggers the activation of thrombocytes and further propagates thrombosis(31, 32) Neutrophil extracellular traps (NETs) are also formed as a result of neutrophil mobilization which likewise triggers an inflammation-triggered thrombosis(33). Regardless of aetiology, the hyperactivation of neutrophils in the acute stage of stroke also enhances neurotoxicity through the production of reactive oxygen species and further promotes the release of enzymes such as collagenase, gelatinase, and heparinase which promotes neurovascular breakdown (34). These pathological changes in neutrophil in the central nervous system are manifested peripherally and are reflected in the degree of change in the NLR.

NLR is an excellent biomarker which clinicians may use because it is readily available and easy to interpret. It provides a handy and inexpensive tool to identify an individual’s state of inflammation, albeit non-specific. Various studies have looked into the utility of NLR in predicting stroke-related mortality, particularly in the short term(35). A linear correlation was demonstrated between NLR values on admission and death after one and three months among patients with ischemic stroke (35). These findings were confirmed in a systematic review that concluded that an NLR of more than five was associated with death after three months among patients with AIS (11). A parallel trend was observed among patients with intracerebral hemorrhage. An observational study including more than 200 patients with spontaneous ICH concluded that an elevated NLR at baseline is likely to result in death after 30 and 90 days(36, 37)

In-hospital mortality for stroke patients, especially in the acute stage is influenced by many factors, including baseline NIHSS, GCS and underlying vascular risk factors(38). Scant information is known as to the potential utility of various biomarkers in predicting mortality, particularly in the acute setting. In particular, biomarkers such as alkaline phosphatase, phosphate, magnesium, BUN and fibrinogen have been shown to have a good correlation within hospital mortality (39-41). This study is the first to date to determine the potential role of elevated NLR in assessing death risk, particularly in the hyper-acute setting.

The value of such an easily available, cheap, immune-based prognostic marker is noteworthy. It is undeniable that families of patients diagnosed with catastrophic stroke are more likely to have difficulty transitioning to end of life care due to the abruptness of the emotional impact (42). Family members of patients with good functionality at baseline are likely to be impacted more. In this study, we have demonstrated that NLR among this subset of patients had a higher value in predicting mortality. This may guide clinicians in communicating prognosis to family members and may smooth transition to end of life care.

This study has limitations. Its retrospective design and the involvement of a small number of patients confined in a single centre may be potential confounders. Combining patients with ischemic and haemorrhagic strokes are also likely to contribute to heterogeneity. Furthermore, using GCS, rather than NIHSS as a crude marker of stroke severity is another limitation.

## Conclusion

The neutrophil to lymphocyte ratio may be utilized as a biomarker for determining the degree of innate and adaptive immune imbalances, most especially in the acute stage of injury. This translates to its potential use as a prognosticating marker for hyper-acute in-hospital mortality, especially in ischemic and haemorrhagic stroke patients.

The NLR is easily available to any clinician who manages these patients anywhere in the world at a very low cost.

Its non-specificity limits its use; however, its practical implications may outweigh its limitations. Systemic immune-inflammation index (SII) (can be easily calculated and likely to add further value towards systems biology approach in prognostication of acute stroke. Serial Systemic immune-inflammation index may provide additional value and further research is warranted as the systems biology based approach is likely to be helpful in diagnosis, progress, prognosis and treatment in the context of post stroke neurological complications.

### Box 1;

**Systemic Immune inflammatory index (SII)**(43, 44)

SII = P x N/L where P, N and L are the cell counts per litre of peripheral blood for platelets, neutrophils and lymphocytes.

## Data Availability

available from the corresponding author with a resonable request

